# Repetitive Transcranial Magnetic Stimulation over Primary Somatosensory Cortex for Upper Limb Function in Stroke: An Exploratory Randomized Controlled Trial

**DOI:** 10.64898/2026.06.15.26355651

**Authors:** Alfredo Lerín-Calvo, Sergio Lerma-Lara, Marcos Moreno-Verdú, Alejandro Herrera-Rojas, Lorena Remón-Ramiro, Carlota López-Tapia, David Rodríguez-Martínez, Raúl Ferrer-Peña

## Abstract

**Background:** Stroke often causes Upper Limb (UL) functional impairments. The Primary Somatosensory Cortex (S1) plays an important role in motor learning. Repetitive Transcranial Magnetic Stimulation (rTMS) over S1 could enhance UL recovery. We aimed to explore its preliminary effects on UL motor activity and function post-stroke.

**Methods:** An exploratory parallel-group randomized controlled trial in people with chronic stroke (>3 months) and moderate hemiparesis was conducted. Participants received 20 sessions of active or sham 5Hz rTMS over affected S1, with Robot-Assisted Therapy and Task-Oriented Training, 5 days/week for 4 weeks. The primary endpoint was UL motor activity (Action Research Arm Test, ARAT). Secondary measures were the UL Fugl-Meyer Assessment (UL-FMA) and sensory outcomes.

**Results:** The baseline-adjusted mean difference (MD) in ARAT was 4.05 points [0.78, 7.33], favoring active stimulation. Secondary measures did not favor active stimulation (UL-FMA: MD = 2.62 [-1.51, 6.76]; sensory outcomes showed no between-group differences).

**Conclusion:** High-frequency rTMS over S1 may enhance UL motor activity (ARAT), but no evidence for motor impairment (UL-FMA) or sensory domains was found. Compensation rather than restoration may underlie this improvement. Stimulation targets should match the intended recovery domain, although larger trials are needed to confirm these preliminary findings.

**Clinical trial registration number:** ClinicalTrials.gov ID: NCT05467657 (https://clinicaltrials.gov/study/NCT05467657?cond=NCT05467657&rank=1)

## INTRODUCTION

Stroke is the second leading cause of mortality and disability worldwide (1), with 45-75% of People with Stroke (PwS) exhibiting Upper Limb (UL) motor impairments (2). Motor recovery relies on two mechanisms (3): restitution, the return towards pre-illness levels of motor control and strength, and compensation, the use of residual effectors, muscles, or joints to achieve a task (4). Restitution occurs predominantly within the first three months post-stroke, whereas compensation predominates thereafter (5).

Interventions focused on improving compensation are encouraged to enhance activities and participation in PwS, such as Task-Oriented Training (TOT) and Constraint-Induced Movement Therapy (6). The main basis of these interventions is motor learning of different activities that must be introduced into individuals’ daily living with behavioral strategies (7,8).

Different brain regions are involved in motor learning, such as the primary somatosensory cortex (S1), primary motor cortex (M1), caudal basal ganglia, and the cerebellum (9). Beyond its traditional sensory role, growing evidence highlights S1 as a critical hub for skill acquisition and sensorimotor integration (9). Animal studies have shown that S1 ablation impairs skill acquisition while enabling consolidation, and disruptive non-invasive brain stimulation impairs motor learning in healthy humans (9,10). Recent evidence supports the role of S1 and other sensory cortices as context_dependent hubs within distributed sensorimotor networks that support detection, decision_making, and learning (11).

Top-down approaches are promising options to enhance UL motor function after stroke by directly modulating cortical networks involved in motor learning and control (12,13). In this context, repetitive Transcranial Magnetic Stimulation (rTMS) is a non-invasive technique that modulates cortical excitability and has been widely applied in neurological and psychiatric disorders (12,14). While rTMS over the primary motor cortex (M1) is established for enhancing restitution (13), the potential of S1 stimulation to support motor control remains underexplored.

Given the significance of S1 in motor learning and the limited capacity for restoration in PwS, we hypothesize that applying high-frequency rTMS to stimulate the lesioned S1 combined with TOT would enhance motor learning in PwS and generate changes in their UL activity. Thus, the aim of this study is to examine the preliminary effects of rTMS over S1 combined with TOT for improving UL motor activity in PwS.

## MATERIALS AND METHODS

### Design and Setting

A parallel-group randomized controlled trial (ID: NCT05467657) was conducted at the Neuron Clinic (Madrid, Spain) and reported following CONSORT criteria (15). It took place between December 2023 and November 2025, and its protocol was published (16). The research adhered to the Declaration of Helsinki (17), and was approved by the “Regional Drug Research Ethics Committee of the Community of Madrid” (internal number: A001). All participants provided written informed consent prior to enrollment.

### Participants and Eligibility Criteria

Consecutive non-probability convenience sampling was performed with participants meeting the following inclusion criteria: 1) first-ever ischemic or hemorrhagic stroke >3 months prior to enrollment; 2) age >18 years; 3) moderate UL motor impairment, scoring 15-53 points (max=66) on the motor subscale of UL Fugl-Meyer Assessment (UL-FMA) (18).

The exclusion criteria were: 1) inability to understand simple sentences or MMSE < 22; 2) any rTMS contraindication (14) (e.g. metallic implants at the cortical level or body-worn electronic devices; history of epileptic seizures or current use of medications that lower the seizure threshold); 3) previous history of any other neurological pathology.

### Randomization & allocation concealment

A researcher not involved in the study used GraphPad software (GraphPad QuickCalcs, USA) to randomly assign the participant to either control (sham rTMS) or experimental (active rTMS) groups in a 1:1 allocation ratio. Allocation was concealed using sealed envelopes.

### Blinding

Evaluators were blinded to participants’ group allocation. Participants were instructed not to disclose details or feelings about their stimulation to the evaluator. Participants were also blind to the allocated treatment (active or sham rTMS protocol, see below). The researcher applying the treatment and the person conducting statistical analyses were not blinded.

### General Procedures

Participants underwent neurophysiological assessments and clinical evaluations at baseline (t0) and post-intervention (t1) with the same blinded evaluator (Fig. 1a). Adverse events were monitored as criteria for study discontinuation.

**Figure 1.**
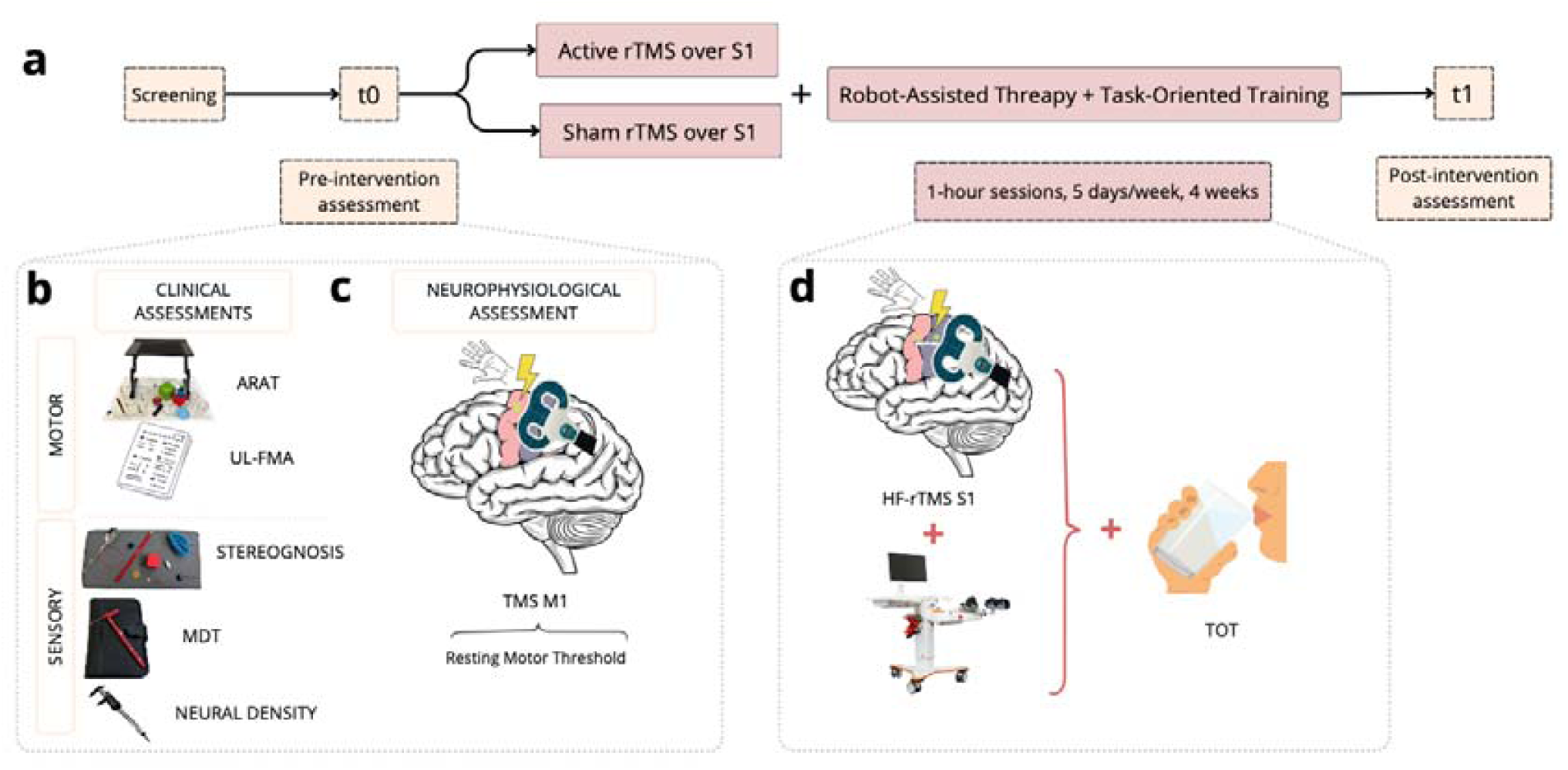
Experiment overview. a) General procedure scheme. b) Clinical Assessments used in the experiment include Action Research Arm Test (ARAT), Upper-Limb Fugl Meyer Assessment (UL-FMA), stereognosis, Mechanical Detection Threshold (MDT) using Semmes-Wenstein monofilaments, and neural density evaluated through the Two Point Discrimination test (TPD). c) Neurophysiological assessment of the Resting Motor Treshold of the PwS with TMS. d) Intervention applied in both groups. Robot-Assisted Therapy (RAT) and Task-Oriented Therapy (TOT) was delivered in all PwS and HF-rTMS over S1 was applied before RAT and TOT for PwS in the experimental group, while sham rTMS was applied in the control group.

### Interventions

Participants completed 20 sessions (5 days/week, 4 weeks)(13) comprising simultaneous rTMS (active or sham) and Robot-Assisted Therapy (RAT), followed by 30 minutes of Task-Oriented Training (TOT) (Fig. 1d).

#### rTMS over S1 intervention

Stimulation was delivered using the STM 9000, ATES MEDICA Device (EBNeuro SpA®, Italy), with a 70mm air-cooled, flat, figure-of-eight coil. Participants sat in a chair with back support in a relaxed posture throughout the TMS procedure.

Daily, the M1 motor hotspot of the First Dorsal Interosseous (FDI) muscle was identified in the non-paretic hand by positioning the coil over the area producing the maximum FDI response (defined as a visible contraction or index finger abduction), with the coil placed tangentially approximately 3cm lateral to the midline with a 45° handle rotation, although the exact position was adjusted individually(19). The Resting Motor Threshold (RMT) was determined as the minimum stimulator output producing visible FDI contraction in ≥5/10 consecutive trials (Fig. 1d) (20).

Every treatment session, the affected M1 was first localized, and the coil was then placed 2cm posterior to the M1 point to find the lesioned S1. To ensure correct positioning, 10 single pulses at 110% of the RMT were applied, confirming the absence of motor responses, as described in previous studies (21). If no motor hotspot could be identified in the affected hemisphere, M1 and S1 were localized by mirroring the coordinates of the intact hemisphere (19).

Participants in the experimental group received the active (real) stimulation protocol (22), consisting of 24 trains of 50 pulses at 5Hz at 90% RMT intensity, inter-train interval of 5 seconds, for a total of 1200 pulses/session. Control group participants received the same stimulation protocol, but once S1 was located, the coil was placed transversely, effectively impeding stimulation, therefore constituting a sham condition (23).

#### Robot-assisted therapy (RAT)

RAT was administered simultaneously with rTMS using the Amadeo® device (Tyromotion), an end-effector robot that generates movement at the distal phalanx of each finger (24). Participants were comfortably seated with armrests, the device height adjusted individually using Velcro straps at the wrist level, and magnets at the distal phalanges facilitated finger positioning.

The protocol consisted of 150 passive finger flexion-extension repetitions followed by 50 active-assisted repetitions, where the robot prompted movement through a screen and provided passive assistance if no movement was detected within 5 seconds, totaling 200 repetitions per session, with movement amplitude, speed, and assistance individually tailored (25).

#### Task-oriented training (TOT)

All participants completed four TOT exercises per week, randomized based on individual UL-FMA subscale scores using a dedicated application (https://github.com/VikinXX/Task-Oriented-Training-Randomizer). Cut-off points and a database with an English version of the tasks used in the randomization are available within the decision-tree flowchart for randomization in the Supplementary Material (SM1, SM2).

Each TOT exercise was performed as many times as possible within 5-minute bouts, up to a total of 30 repetitions, based on the foundational principles of repetitive task training interventions (26). Once they reached 30 repetitions for an exercise, it was made harder to ensure progression in task requirements (27). If participants were unable to perform any repetitions of an exercise within the 5 minutes, they were asked if they wanted to continue trying the same exercise the next day or if they would prefer to make the task easier or change to an easier exercise to avoid frustration and excessive task difficulty(26). Independent of the weekly results, exercises were changed every Monday. Participants’ progression during the intervention period was registered to ensure transparency and is available in the Supplementary Material (SM3).

### Outcome measures

The primary outcome of the study was UL motor activity (28) using the Action Research Arm Test (ARAT) (Fig. 1b). This 19-item tool is a quantitative test with four subtests: grasping, holding, pinching and gross motor skills (29). Each item is scored from 0 to 3: 0 = unable to perform the task; 1 = can perform part of the task; 2 = completes the task but takes an excessively long time or great difficulty; 3 = performs the task normally. The total score ranges from 0-57 points, higher scores indicating better performance.

UL motor function impairment, RMT, and various sensory domains were assessed as secondary outcomes. Corticospinal excitability was assessed via RMT(20).

UL motor function impairment(4), was assessed using the UL-FMA. This test measures the ability to move different joints out of synergies, with scores ranging from 0-66 points, higher scores reflecting better motor function (30,31).

Sensory domains included

- Stereognosis, defined as the three-dimensional perception and recognition of an object through active touch, was evaluated by asking participants to identify a series of objects presented randomly between their fingers and thumb, with their eyes closed, from the ulnar side of their hand. The test objects included a pencil, a paper clip, a safety pin, a coin, a button, a pill, a rubber band, a string, a spoon, a cube, and a marble (32). The score ranged from 0-11 points, higher scores indicating better performance.
- The Mechanical Detection Threshold (MDT) was evaluated using Semmes-Wenstein monofilaments of varying weights, applied to a specific point on the thenar eminence. The stimulus with the heaviest weight, 4.5g/mm, was presented to the participant with their eyes closed. If the participant felt it three times, the next monofilament, with a lower weight, was applied until the participant could no longer recognize the stimulus. This process was repeated three times to obtain an average (33). The score was provided in millimeters, higher scores indicating better performance.
- Neural density was assessed using the two-point discrimination test (TPD). Using a caliper, single- or two-point touches were applied to the thenar eminence of the affected hand in sets of three stimuli. The distance started at 50mm and was decreased by 5mm each time until all three stimuli were correctly selected. If a stimulus was missed, the result was recorded (34). The score was provided in centimeters, lower scores indicating better performance.

### Sample Size Calculation

A priori sample size was estimated following the accuracy in parameter estimation approach (35) in R v4.2. Using a previous study (36), the ARAT’s SD was set at 12 points in both groups. As no prior study reported the repeated-measures correlation for the ARAT, a conservative baseline–post-intervention correlation of r=0.50 was assumed. The target confidence-interval half-width was fixed at ±6 points (95% confidence, 80% assurance), with 10% anticipated attrition, yielding a planned N=64 (32 per group). Recruitment was stopped prematurely at N=20 owing to resource constraints, before the planned sample was reached; the trial is therefore underpowered, and all analyses are interpreted as exploratory.

### Data Analysis

All analyses were run in R v4.5.1 (R Core Team 2026). For clinical scales with inherent lower and upper bounds (e.g. ARAT, UL-FMA, stereognosis), beta-regression models were fitted using package ‘glmmTMB’(37,38). Scores were first rescaled to 0-1 before fitting the model. Model-based predictions and comparisons are reported back-transformed to the original scale.

Continuous outcome measures (e.g., MPT, TPD) did not follow a normal distribution and showed evidence of heteroskedasticity, and therefore robust linear regression models were fitted using the package ‘MASS’ (39,40). These models were fitted by iterated re-weighted least squares via M-estimation. Uncertainty around predictions and comparisons was computed by heteroskedasticity-consistent standard errors (HC3) (41).

Inference was based on ANCOVA-type models (including the baseline measure as a covariate) with the formula: Post ∼ Pre + Treatment. Post-hoc tests between active and sham groups were asymptotic Wald z-tests using package ‘marginaleffects’ (42). The average treatment effect was the post-intervention, baseline-adjusted, between-group mean difference (43). For statistical significance we used alpha=5%.

## RESULTS

Overall, 20 participants were included, randomly assigned, received the interventions as intended and attended the assessment sessions, without dropouts or missing data (Fig. 2). No adverse events were reported in any group. Table 1 shows baseline data.

**Figure 2.**
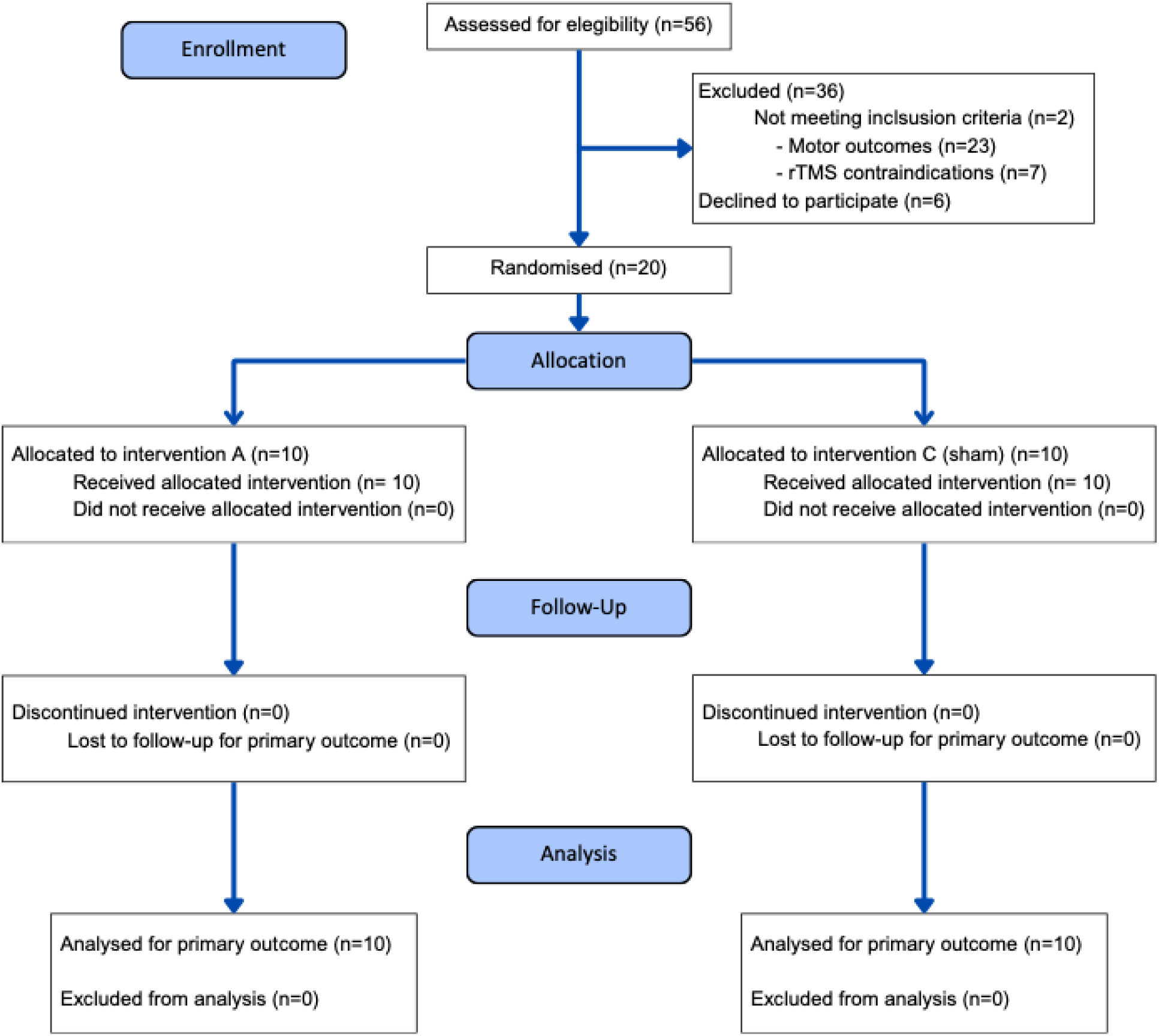
Flowchart diagram of the participants in the study.

**Table 1.** Participants’ characteristics at baseline.

| Characteristic | Active<br>N = 10<sup>1</sup> | Sham<br>N = 10<sup>1</sup> | Overall<br>N = 20<sup>1</sup> |
| --- | --- | --- | --- |
| Sex |  |  |  |
| Female | 2 (20%) | 1 (10%) | 3 (15%) |
| Male | 8 (80%) | 9 (90%) | 17 (85%) |
| Age (years) | 44.00 (10.01) | 52.10 (16.76) | 48.05 (14.06) |
| Education |  |  |  |
| College graduate or more | 6 (60%) | 3 (30%) | 9 (45%) |
| High school graduate | 1 (10%) | 5 (50%) | 6 (30%) |
| Middle school graduate or less | 3 (30%) | 2 (20%) | 5 (25%) |
| Time since stroke (months) | 30.90 (58.06) | 15.90 (10.47) | 23.40 (41.33) |
| Type of stroke |  |  |  |
| Hemorrhagic | 3 (30%) | 6 (60%) | 9 (45%) |
| Ischemic | 7 (70%) | 4 (40%) | 11 (55%) |
| Affected hemisphere |  |  |  |
| Left | 4 (40%) | 5 (50%) | 9 (45%) |
| Right | 6 (60%) | 5 (50%) | 11 (55%) |
| Action Research Arm Test (0-57 points) | 14.40 (17.49) | 5.70 (8.76) | 10.05 (14.18) |
| Fugl-Meyer Assessment-Upper Limb (0-66 points) | 30.20 (11.53) | 22.10 (4.68) | 26.15 (9.52) |
| Stereognosis (0-11 points) | 3.30 (3.23) | 1.80 (2.70) | 2.55 (3.00) |
| Mechanical Pressure Threshold (kg) | 0.95 (1.33) | 2.65 (1.68) | 1.80 (1.71) |
| Two-point discrimination (mm) | 40.30 (23.66) | 35.50 (28.72) | 37.90 (25.73) |
| Presence of MEP (affected hemisphere) |  |  |  |
| MEP- | 3 (30%) | 7 (70%) | 10 (50%) |
| MEP+ | 7 (70%) | 3 (30%) | 10 (50%) |
| RMT - Affected hemisphere (% MSO) | 63.57 (16.95) | 59.00 (9.54) | 62.20 (14.72) |
| Not found | 3 | 7 | 10 |
| RMT - Unaffected hemisphere (% MSO) | 52.70 (8.94) | 57.80 (15.11) | 55.25 (12.37) |

<sup>1</sup>Values are presented as mean (SD) or n (%).
<sup>1</sup>Abbreviations: MEP: Motor Evoked Potential; RMT: Resting Motor Threshold; % MSO: Percentage of Maximal Stimulator Output.

For ARAT, the average treatment effect at post-intervention favored the experimental group (mean difference = 4.05 points [0.78, 7.33], z = 2.43, p = 0.0153) (Fig 3.). For UL-FMA, no differences were found between groups (mean difference = 2.62 points [-1.51, 6.76], z = 1.25, p = 0.2131) (Fig. 4a). No differences were found between groups for any of the sensory outcomes, including stereognosis (mean difference = 0.12 points [-1.05, 1.29], z = 0.2, p = 0.8433) (Fig. 4b), MPT (mean difference = −0.37 kg [-1.53, 0.8], z = −0.62, p = 0.5383) (Fig. 4c), and TPD (mean difference = −9.02 mm [-20.83, 2.78], z = −1.5, p = 0.134) (Fig. 4d). No results could be obtained from RMT due to lack of motor hotspot in most participants (see Table 1).

**Figure 3.**
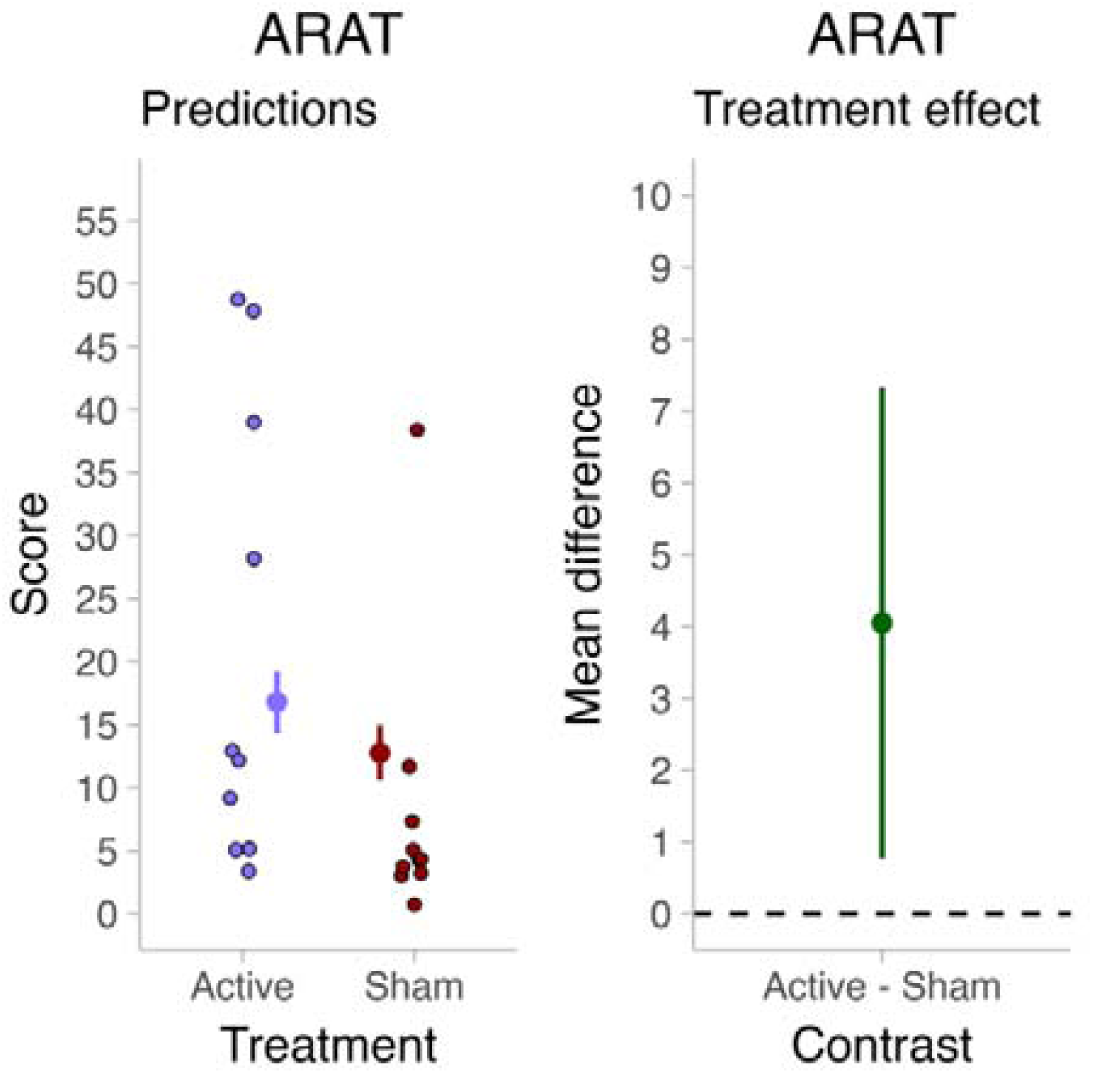
Results of the primary end-point, Action Research Arm Test (ARAT) at post-intervention. Panel **a)** shows model-predicted (baseline-adjusted) marginal means for the experimental (active rTMS) or control (sham rTMS) groups. Panel **b)** shows the average treatment effect (mean difference between groups).

**Figure 4.**
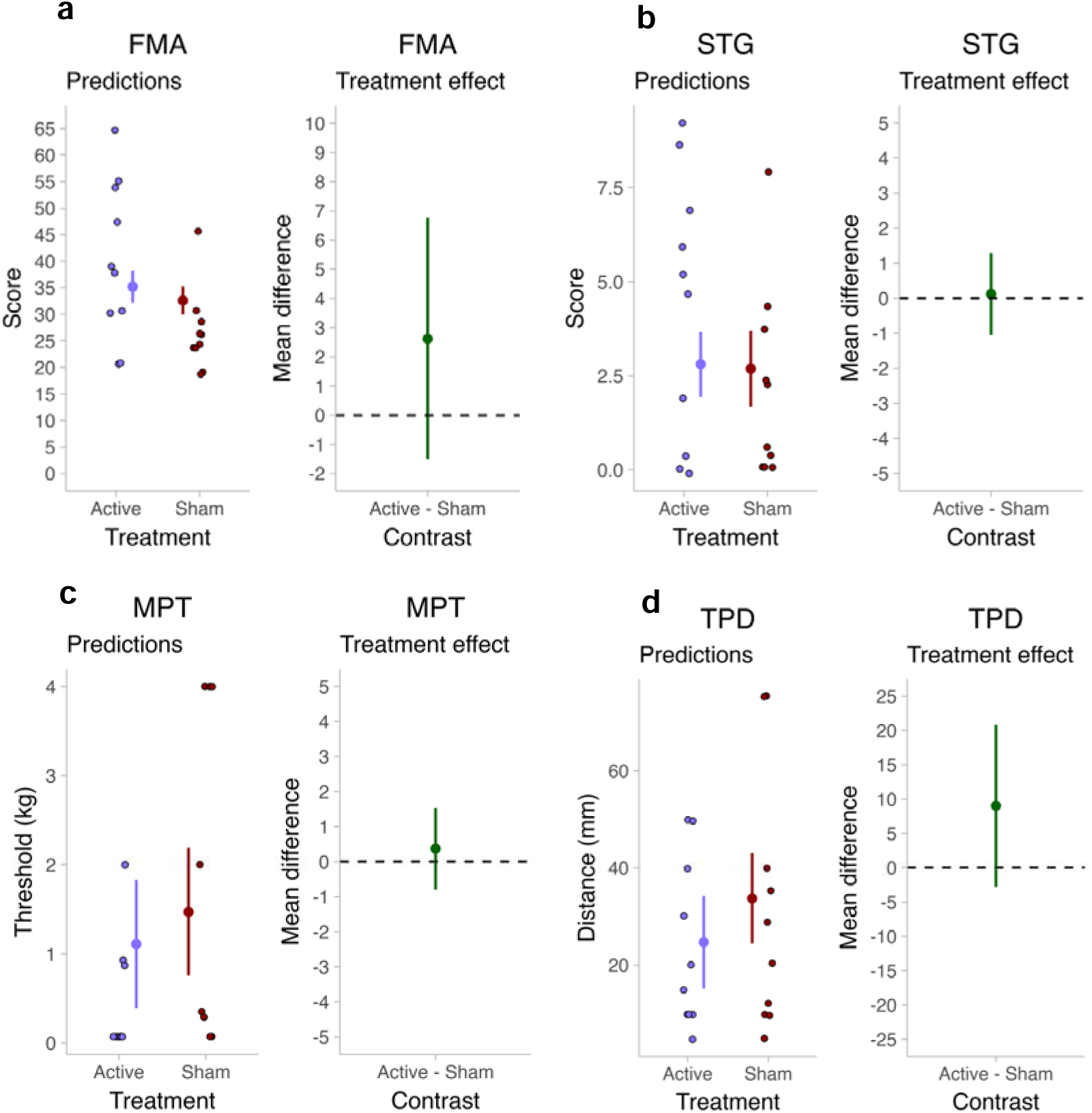
Results of the secondary end-points, including the UL Fugl-Meyer Assessment (UL-FMA), Stereognosis (STG), Mechanical Pressure Threshold (MPT) and Two-point Discrimination (TPD), at post-intervention. On each panel, the plot on the left shows model-predicted (baseline-adjusted) marginal means for the experimental (active rTMS) or control (sham rTMS) groups. The plot on the right shows the average treatment effect (mean difference between groups).

## DISCUSSION

We explored the preliminary effects of high-frequency rTMS over S1 combined with RAT and TOT in improving UL motor activity in PwS. We found that the intervention enhanced UL motor activity compared to sham stimulation, with a baseline-adjusted mean difference of 4.05 points in ARAT. However, no significant differences were found for motor control impairment (UL-FMA) or sensory outcomes (stereognosis, mechanical pressure threshold, two-point discrimination). These results suggest that S1 stimulation primarily facilitates compensatory motor learning and functional task performance rather than restitution of underlying motor impairments.

### UL activity

The most targeted area for improving UL motor function in PwS is M1 (44). However, recent meta-analyses suggest that it has no effect on UL activity, but can improve motor control and corticospinal excitability (45). Our results demonstrate that S1 stimulation combined with motor training improved UL activity. Although the effect size is modest, it is particularly relevant considering the chronic stage of our participants and the limited potential for spontaneous recovery beyond the first three months post-stroke (46).

To our knowledge, there are few studies applying rTMS over S1 in PwS. A proof-of-principle study in chronic stroke reported that rTMS delivered to contralesional S1 can modulate UL somatosensory function, supporting the feasibility and biological plausibility of S1 as a therapeutic target (47). The only previous RCT using 5Hz rTMS over ipsilesional S1 found no between-group differences in motor learning when assessed with a Serial Reaction Time Task (SRTT), which primarily engages cortico-striatal procedural memory systems (22). This discrepancy likely reflects the different systems each task engages: the SRTT is an implicit sequence-learning task relying on cortico-striatal loops and procedural memory (48), whereas dexterity tasks, such as those measured by UL activity tests (eg: ARAT), require explicit motor planning, sensory-guided manipulation, and adaptive control that depend critically on cortical S1–M1 integration (11). Our results are consistent with findings in healthy individuals showing that S1 plays a more prominent role in use-dependent motor learning and tasks requiring sensorimotor integration than in purely sequential or habitual learning (49).

While both M1 and S1 are involved in the motor learning process (10), S1 seems to play a major role when learning new movements (50). Furthermore, recent evidence from human neuroimaging and electrophysiological studies supports the role of S1 as a context-dependent hub within distributed sensorimotor networks that support detection, decision-making, and learning (51). Plastic changes in S1 have been shown to predict motor learning success, while disrupting S1 activity blocks motor memory consolidation (52). Thus, our results suggest that S1 stimulation may be particularly effective for enhancing motor learning in tasks that emphasize sensorimotor integration and functional dexterity.

### Motor control

An important finding in our study is the dissociation between improvements in UL activity (ARAT) and the absence of changes in motor control impairment (UL-FMA). These instruments assess different constructs within the ICF framework: the ARAT measures activity (i.e., the ability to perform functional tasks in standardized conditions), whereas the UL-FMA quantifies impairment (i.e., the capacity to move individual joints in isolation, independent of synergistic patterns) (53). This distinction is critical for understanding the mechanisms underlying motor recovery after stroke.

Our results suggest that S1 stimulation primarily facilitated compensatory motor learning, improving the ability to perform functional tasks through refined sensorimotor integration and optimized movement strategies, rather than restitution of impaired motor control. The lack of change in the UL-FMA suggests that S1 stimulation did not significantly alter the underlying impairment in selective motor control, reduce synergistic movement patterns, or enhance corticospinal excitability.

Unfortunately, RMT could not be analyzed as a secondary outcome because 50% of participants lacked reliable motor-evoked potentials (MEPs) in the affected hemisphere, reflecting the degree of corticospinal tract damage common in chronic stroke populations with moderate-to-severe impairment (54). The presence of MEPs has been shown to be a strong predictor of motor recovery potential, and PwS without MEPs typically have more severe corticospinal tract damage and rely more heavily on compensatory mechanisms for functional recovery (55).

These results contrast with the effects typically observed with high-frequency M1 stimulation, which increases corticospinal excitability and can improve UL-FMA scores, reflecting restitution-related changes, although this comparison must be interpreted cautiously given our small sample (44,45). Nonetheless, the potential limitations of UL-FMA as a measure of motor control can conflate genuine improvements in selective motor control with increases in strength (56). Some authors argue that the UL-FMA may not fully capture the quality of movement or adequately distinguish between true restitution and compensation, as PwS may learn to perform the test items more efficiently without necessarily recovering normal motor control (5). Our results could suggest that PwS improved UL-FMA due to non-specific effects of intensive motor practice more related with strength gains rather than specific changes in corticospinal integrity or motor map organization (57).

Our findings support the broader principle that different brain stimulation targets may be optimal for different aspects of motor recovery and different phases of rehabilitation. This need for personalization is consistent with recent high-quality syntheses showing that rTMS effects on motor outcomes vary with stimulation type, target site, and pathology stage, underscoring that target selection (e.g., M1 vs S1) should be matched to the intended recovery domain (impairment vs activity) and individual neurobiological reserve (58).

### Sensory domains

We hypothesized that high-frequency rTMS over S1 would enhance somatosensory function, given S1’s primary role in processing tactile, proprioceptive, and discriminative sensory information (59). However, no significant between-group differences were found for any sensory outcome.

Our results are opposite to those obtained in a previous study which observed an effect in TPD (22). This can be explained by the motor training applied in our study through RAT and TOT, because both interventions inherently involve sensory feedback and haptic stimulation, which may have provided sensory training to both groups, consistent with evidence that adding a specific sensory intervention to task practice does not further improve sensory outcomes (60).

Recent preliminary evidence also suggests that the direction of S1 neuromodulation may matter for somatosensory outcomes. A randomized trial in acute/subacute stroke compared excitatory versus inhibitory rTMS over S1 and reported differential effects on somatosensory functioning, indicating that a ‘one-size-fits-all’ excitatory protocol may not uniformly translate into measurable improvements on standard clinical sensory tests across recovery stages. This aligns with the possibility that, in chronic stroke and in the presence of intensive haptic-rich training in both groups (RAT/TOT), any incremental sensory benefit of excitatory S1 stimulation could be small and difficult to detect with coarse threshold-based measures (61).

Furthermore, the sensory outcome measures used may lack sensitivity to detect subtle cortical-level changes. Stereognosis, for example, is a complex, high-level sensory task that depends not only on intact tactile discrimination and proprioception but also on cognitive factors (59). Similarly, TPD and MDT assess basic sensory detection capacities that are closely tied to peripheral receptor density and early sensory processing, and these measures may be less responsive to modulation by cortical stimulation, particularly in the context of chronic sensory deficits with established thalamocortical reorganization (62). More sensitive measures, such as tactile temporal discrimination or sensory-evoked potentials, may better capture the effects of S1 modulation on sensory outcomes (63).

### Strengths, Limitations and Future Directions

This constitutes the initial research evaluating the impact of rTMS over S1 on motor control in PwS, combining rTMS with RAT and TOT. Outcome measures cover multiple ICF domains, including motor activity (ARAT), motor control (FMA-UL) and somatosensory function, providing a comprehensive picture of sensorimotor changes, However, this study was not designed to confirm efficacy but to generate hypotheses and inform the design of a future adequately powered trial

The small number of participants is the major limitation to extrapolating our results. In addition, neuronavigation was not available to confirm precise S1 targeting and to ensure fully consistent coil positioning across all sessions, and the absence or instability of MEPs in some participants may have affected the reliability of RMT-based dosing. Treatment therapists were unblinded to group allocation, which could introduce performance bias despite standardized protocols, and no dedicated sham coil allowing full blinding of treatment administration was used. Furthermore, no long-term follow-up (e.g., 3 or 6 months) was performed, precluding conclusions about the sustainability of the observed improvements over time.

Future research incorporating head-to-head comparisons with M1 stimulation, comprehensive neurophysiological and neuroimaging assessments, longer follow-up periods, and more diverse populations will be essential for establishing the clinical utility of S1 stimulation.

## CONCLUSION

In summary, this RCT provides preliminary evidence that high-frequency rTMS over S1 may produce modest improvements in UL motor activity in chronic stroke with moderate hemiparesis, without significant effects on motor impairment or sensory function. These results support the hypothesis that improvements in UL activity involve enhancement of compensatory strategies and sensorimotor integration rather than restitution of underlying impairments. The dissociation between ARAT and UL-FMA outcomes reinforces the importance of selecting stimulation targets aligned with the intended recovery domain.

## ACKNOWLEDGEMENTS

We express our sincere gratitude to Neuron Clinics for providing the facilities and acquiring the transcranial magnetic stimulator used in this study. We especially thank Clementa Mota Valbuena and Claudia Moreno Rugama for their support and assistance with the participants’ schedule and Álvaro Ortiz Villarejo for its help creating the randomizer app. We also thank the Professional Association of Physiotherapists of the Community of Madrid for supporting research projects and scientific development, which funded this article. Finally, we are deeply grateful to all the participants in the study for their time, commitment, and valuable contributions to the research.

## DATA AVAILABILITY

Databases and scripts used in the study’s statistical analysis are freely available at: https://doi.org/10.5281/zenodo.18633300

