## Supplementary material for "Repetitive Transcranial Magnetic Stimulation over Primary Somatosensory Cortex for Upper Limb Function in Stroke: An Exploratory Randomized Controlled Trial": SM1, SM2

**SM1**. Cut-off points and activities for Task-Oriented Training.

The table below reflects the activities that were randomized for the patient based on its Upper-Limb Fugl Meyer (UL-FMA) scores.

Activities in the upper limb column were always randomized for all the participants, however, the activities in other columns were only included in the randomization if:

- For wrist activities, the participant should obtain more than 4 points in the wrist subscale of the UL-FMA.
- For hand activities, the participant should obtain more than 6 points in the hand subscale of the UL-FMA.
- For wrist activities, the participant should obtain more than 2 points in the coordination subscale of the UL-FMA.

| **UPPER LIMB** | **WRIST** | **HAND** | **COORDINATION / SPEED** |
| --- | --- | --- | --- |
| Turn on/off the light | Set the table for eating | Flip the cards | Bounce a ball |
| Raise/Lower toilet lid | Take out/Put away cutlery | Fold and place napkins | Catch a moving marble |
| Open/Close bathroom door | Take out/Put away dishes from cabinet | Put clothespins on clothes | Return ball with ping pong paddle |
| Drink water | Eat with a spoon (solids) | Remove clothespins from clothes | Move ball from one side to the other |
| Open/Close cabinet | Cut putty | Rearrange clothes on drying rack |  |
| Open/Close drawers | Pour water from one container to another | Spoon coffee into a container |  |
| Pull out drawers | Arrange the bathroom shelf | Dexterity puzzle |  |
| Pull papers from the dispenser | Brush teeth | Turn pages of a book I and II, III, IV, V |  |
| Table cleaning | Make the bed | Flip domino tiles |  |
| Wash hands | Put the pillow in the pillowcase | Carry marbles between containers with fingers I and II, III, IV, V |  |
| Remove suctions from glass | Fold clothes and carry from bed to table | Pick chickpeas from a basket with rice |  |
| Move container from one side of the table to the other | Organize the shelf | Open/Close with the key |  |
| Place objects on the shelf | Organize the wardrobe | Flip Jenga tiles (4x2) |  |
| Hang clothes on the rack | Transfer objects between containers using tongs | Deal cards for 4 people |  |
| Take down clothes from the rack | Place and adjust pillow without using body | Put headband on with both hands |  |
| Clean the glass | Remove pillowcase | Carry large jar up and down |  |
| Wash the dishes | Place gummy (thera flex) on the shelf | Remove clips from the frame |  |
| Open/Close fridge | Carry deodorant to armpits with | Grab marbles from container with all fingers |  |
| Raise/Lower faucet | Take out grey bucket from above | Pick marbles and carry them from tray at 90 degrees |  |
| Place toilet paper |  | Carry blue and yellow pieces upward |  |
| Transport plastic glass (high surface) |  | Move blue pieces from one place to another |  |
| Put in/Take out plastic plate from the fridge |  | Insert key and turn |  |
| Transfer clothes basket |  | Carry hangers to rack |  |
| Transport folded clothes in basket one by one |  | "Rubber nail polish" |  |
| Move glass from table to drawer |  |  |  |
| Carry a glass of water to upper platform |  |  |  |
| Hang clothes in the wardrobe |  |  |  |
| Hang/Unhang clothes from wardrobe |  |  |  |
| Hang clothes on hanger and put on coat rack |  |  |  |
| Pull the jacket |  |  |  |
| Move objects from wardrobe to below |  |  |  |
| Take jacket off the hanger |  |  |  |

**SM2**. Decision-tree Flowchart for TOT activities

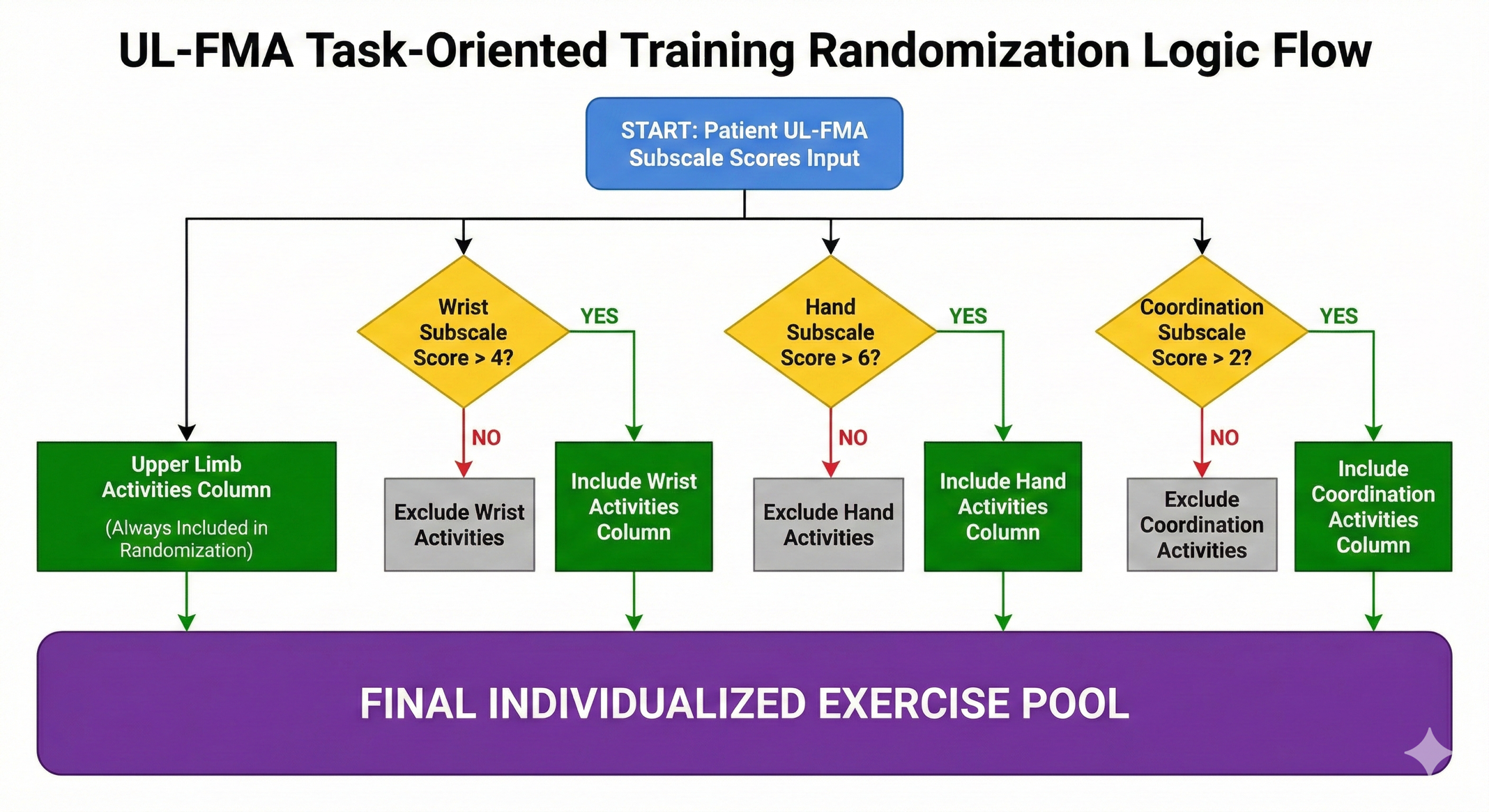
